# Is Covid-19 Mortality “Like the Flu”? A Cumulative Death Rates Comparison

**DOI:** 10.1101/2023.04.24.23289045

**Authors:** Elizabeth Wrigley-Field, Jessie Himmelstern

## Abstract

It has been common both to make and to resist comparisons that equate the Covid-19 pandemic to influenza. We take the comparison between Covid-19 and flu seriously by asking how many years of influenza and pneumonia deaths are needed for cumulative deaths to those two causes to equal the cumulative toll of the Covid-19 pandemic between March 2020 and February 2023—that is, three years of pandemic deaths. We find that in one state alone—Hawaii—three years of Covid-19 mortality is equivalent to influenza and pneumonia mortality in the three years preceding the Covid-19 pandemic. For all other states, at least nine years of flu and pneumonia are needed to match Covid-19; for the United States as a whole, seventeen years are needed; and for four states, more than 21 years (the maximum observable) are needed. These results provide an easy-to-understand calibration of flu as a heuristic for Covid-19, and vice versa.

## MAIN TEXT

In debates about Covid-19 mitigation strategies, it has been common both to make and to resist comparisons that equate the pandemic to influenza (Petersen 2021). Using flu as a yardstick makes sense: making complex and nuanced risk assessments is cognitively challenging, and often relies on interpreting population-level statistics whose implications are highly abstract from the perspective of one individual life. Thus, in many different contexts, people rely on heuristics—like comparing novel risks to familiar ones, for which behavioral repertoires are already established—to gauge risks whose magnitudes are difficult to conceptualize directly (Raue and Scholl 2018). Such heuristics do not arise neatly from intrinsic features of a health risk; they also reflect the socio-political processes that frame a particular disease as more or less risky and more or less important to mitigate (Shiffman and Shawar 2022). When the comparison to flu is used in the context of a larger argument against Covid-19 mitigation, it reflects a context in which, in the United States, flu is socially conceptualized as a minor risk (American Heart Association 2022) despite causing tens of thousands of deaths annually. This conceptualization is underscored by linguistic practice: “flu” is informally used as a designator for a wide variety of minor ailments (including unrelated “stomach flus” and the widespread, but highly heterogeneous, category “colds and flus”). In a country where one in four to one in five workers lack paid sick leave (Pew Research Center 2020; United States Bureau of Labor Statistics 2021) and others, who have it, are expected not to use it, minimizing the potential severity of respiratory illness may have a particular cultural logic.

In this data visualization, we take the comparison between Covid-19 and flu seriously by asking how many years of influenza and pneumonia deaths are needed for cumulative deaths to those two causes to equal the cumulative toll of the Covid-19 pandemic between March 2020 and February 2023—that is, three years of pandemic deaths. We group influenza and pneumonia together because it is common for pneumonia to be the ultimate cause of death following influenza infection. We use data from the National Center for Health Statistics and include as “Covid-19 deaths” only deaths where Covid-19 is assigned as the underlying—not merely a contributing— cause of death. (An alternative analysis using all deaths involving reported Covid-19 is shown in the Appendix.) Influenza and pneumonia deaths are also those in which either one was listed as the underlying cause of death. Because deaths are reported on a lag—death reporting is typically about 75% complete eight weeks after deaths occur (Spencer and Ahmad 2023)—the Covid-19 cumulative death rates, and hence the time needed for influenza and pneumonia death rates to match them, will be slightly underreported.

In calculating the influenza and pneumonia seasons needed to match deaths from the Covid-19 pandemic, we begin at the end of 2019 (just before the start of the Covid-19 pandemic in the United States) and move backward in time, in units of one year, until cumulative mortality rates to influenza and pneumonia equal cumulative Covid-19 mortality rates. We avoid incorporating flu and pneumonia deaths during the Covid-19 pandemic because the pandemic substantially altered flu mortality. The 2020-2021 flu season was minimal due to social distancing (Go and Elango 2022; Stojanovic et al. 2021). Moreover, there has been persistent, and still unresolved, debate about how many deaths attributed to influenza or pneumonia—but not Covid-19—were actually due to Covid-19 (Faust and del Rio 2020; Zhu, Ozaki, and Virani 2021). We sidestep those questions by beginning our backwards-running clock just before the Covid-19 pandemic. Finally, we calculate the time needed to equalize cumulative Covid-19 death rates—not cumulative Covid-19 deaths—so that the comparison is not distorted by population growth during the period we study. Further methodological details and some additional graphs are in the Appendix.

Results are shown in Figure 1. For the United States as a whole, the Covid-19 pandemic through February 2023 was the equivalent of 17 years of flu and pneumonia mortality. There is substantial variation in this outcome across states. Hawaii needs only 3 years: its flu and pneumonia death rates from 2017-2019 exceed its Covid-19 death rates from March 2020 to February 2023 (also a three-year span). Hawaii excepted, states range from 9 to more than 21 years (the maximum time period covered in the available data), with a median of 16 years. The results in general show substantial variation across states which is not associated in any obvious way with geographic region, state politics, or state demographics, perhaps reflecting in part that some risk factors that drive heavy Covid-19 mortality have also driven flu and pneumonia mortality for a long time. Variation notwithstanding, the minimum length of time to match Covid-19 mortality, with the exception of Hawaii, is three times longer than the Covid-19 period analyzed here, and usually much longer.

**Figure 1.**
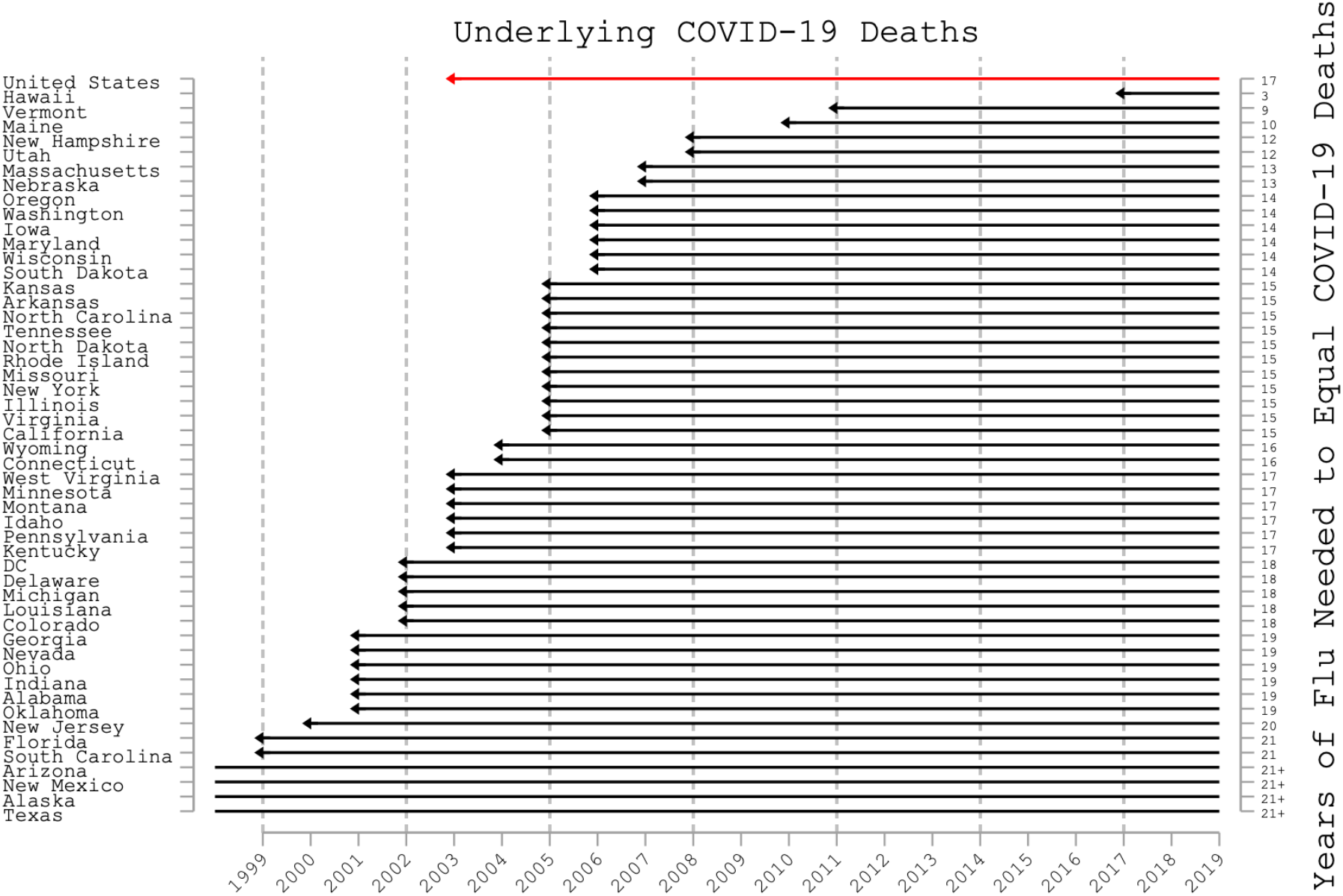
Years required for death rates to influenza and pneumonia (beginning in 2019 and counting backwards) to match the cumulative death rate to Covid-19 over a three-year period (March 2020-February 2023), for the United States as a whole (top line) and by state. The Covid-19 deaths included are those in which Covid-19 is listed as the underlying cause of death, and likewise for influenza and pneumonia.

Hawaii’s exceptional outcome is instructive. In Hawaii, alone, the Covid-19 pandemic is “like the flu.” This reflects a relatively unusual flu history in Hawaii in which influenza and pneumonia deaths have, in general, risen steadily since 2008 (see Appendix Figure A1) and in which 2017 was an exceptionally harsh flu year (with 11 of 2017’s 12 months having either the highest or second-highest rate of flu and pneumonia deaths in Hawaii for that month in 2009-2019). Thus, counting backwards, we find that Hawaii needs go only back to 2017 to match Covid’s cumulative death rate.

Figure 1 supports the perspective that the Covid-19 pandemic has had consequences quite unlike influenza and pneumonia. It takes 17 years for the United States to accrue as much cumulative mortality to these causes as were accumulated to Covid-19 in three years. For four states, this comparison outstrips the available data: one would have to amass deaths from 2019 to even earlier than 1999 to match Covid-19 death rates. On the other hand, even as it highlights the drastic difference between flu and Covid-19, Figure 1 can also be read to underscore the lethality of flu. Covid-19 is a once-a-century pandemic that has led to historic drops in life expectancy all over the world, including in the United States for consecutive years (Arias et al. 2022). The 17 years of flu and pneumonia required to match this cumulative rate of death is more than five times longer than Covid-19 took to enact the same toll—but flu is not once-a-century phenomenon. From that perspective, Figure 1 underscores the vast toll of respiratory viruses broadly for health and survival in the United States. In that sense, these results not only provide an easy-to-understand calibration of influenza as a heuristic for the scale of Covid-19 mortality; they also provide calibration for the converse heuristic as well.

## Data Availability

All data and statistical code are available at https://doi.org/10.17605/OSF.IO/5JS8B

## ACKNOWLEDGEMENTS

The authors gratefully acknowledge helpful assistance from Kriti Budhiraja and Michelle Niemann. Both authors received support from the Eunice Kennedy Shriver National Institute for Child Health and Human Development via the Minnesota Population Center (P2C HD041023).

## APPENDIX TO

In this appendix, we elaborate on the construction of the measures and provide some additional results.

Death rates are constructed using death counts and population estimates from the National Center for Health Statistics (NCHS), as accessed through CDC Wonder. All data and code are available in <u>our public data repository</u>. Covid-19 death counts were downloaded on March 17, 2023 which implies that deaths in 2023 will be undercounted. However, since death rates in the months leading up to the data download were low, the undercounting is not likely to affect cumulative death rates very much.

We use annual death counts in order to avoid extensive missing data problems that arise with smaller time units; the NCHS suppresses death counts for any cells in which there are fewer than 10 deaths, which characterizes many months of influenza and pneumonia deaths in small states or in months outside of flu season. For the same reason, we also use calendar years rather than years defined by flu seasons (whose timing is variable but is typically, roughly, October-March); only calendar years are reported directly as annual units in CDC Wonder, minimizing data suppression.

Population counts are reported by the Census Bureau and NCHS for 1999-2021; we extend them through 2023 using log-linear extrapolation. Estimated population sizes from these agencies use the previous decadal Census as a “base” and extend the population forward in time from that base using the annual American Community Survey alongside some demographic modeling. For 2020, two sets of estimates are available, one using the 2010 Census base (used for 2010-2020) and one using the 2020 Census base (used for 2020-2021). Ordinarily, the more recent base would be preferable, but due to concerns about the quality of the 2020 Census, we average the two estimates for 2020 to produce a unified series without a sharp break in either 2020 or 2021.

Because population sizes have increased over the period we study, our analysis equalizes death rates rather than counts of death. Our algorithm recalculates cumulative death rates for each period moving backward from 2019, which implicitly population-weights the cumulative rates by pooling across periods.

In order to explore Hawaii as the outlier state in which our analysis shows Covid-19 deaths as being “like the flu,” Figure A1 shows Covid-19 and influenza and pneumonia death rates in the relevant years in Hawaii only. Figure A1 reveals the large increase in influenza and pneumonia death rates from 2012 onward, as well as the large spike in 2017, producing this result.

Figure A2 shows an alternative version of our main Figure 1, using Covid-19 deaths that were listed anywhere on a death certificate instead of only as the “underlying” cause of death; in other words, deaths that list Covid-19 as a “contributing” cause, which were excluded from the main analysis, are included here. The historical cause of death reporting does not allow us to also include deaths, for 1999-2019, in which influenza and pneumonia were contributing causes, so they are only included when underlying causes. For this reason, Figure 1 is preferred, but Figure A2 offers a point of comparison. Figure A2 also includes state lines colored by region, which reveals that the states in which it would take the fewest years for flu and pneumonia deaths to match Covid-19 deaths are concentrated in the Northeast and West, although northeastern and western states are also scattered across the range of this outcome variable. Moreover, none of the thirteen states in which the fewest flu and pneumonia deaths would be required to match cumulative Covid-19 deaths are in the South.

### APPENDIX FIGURES

**Figure A1.**
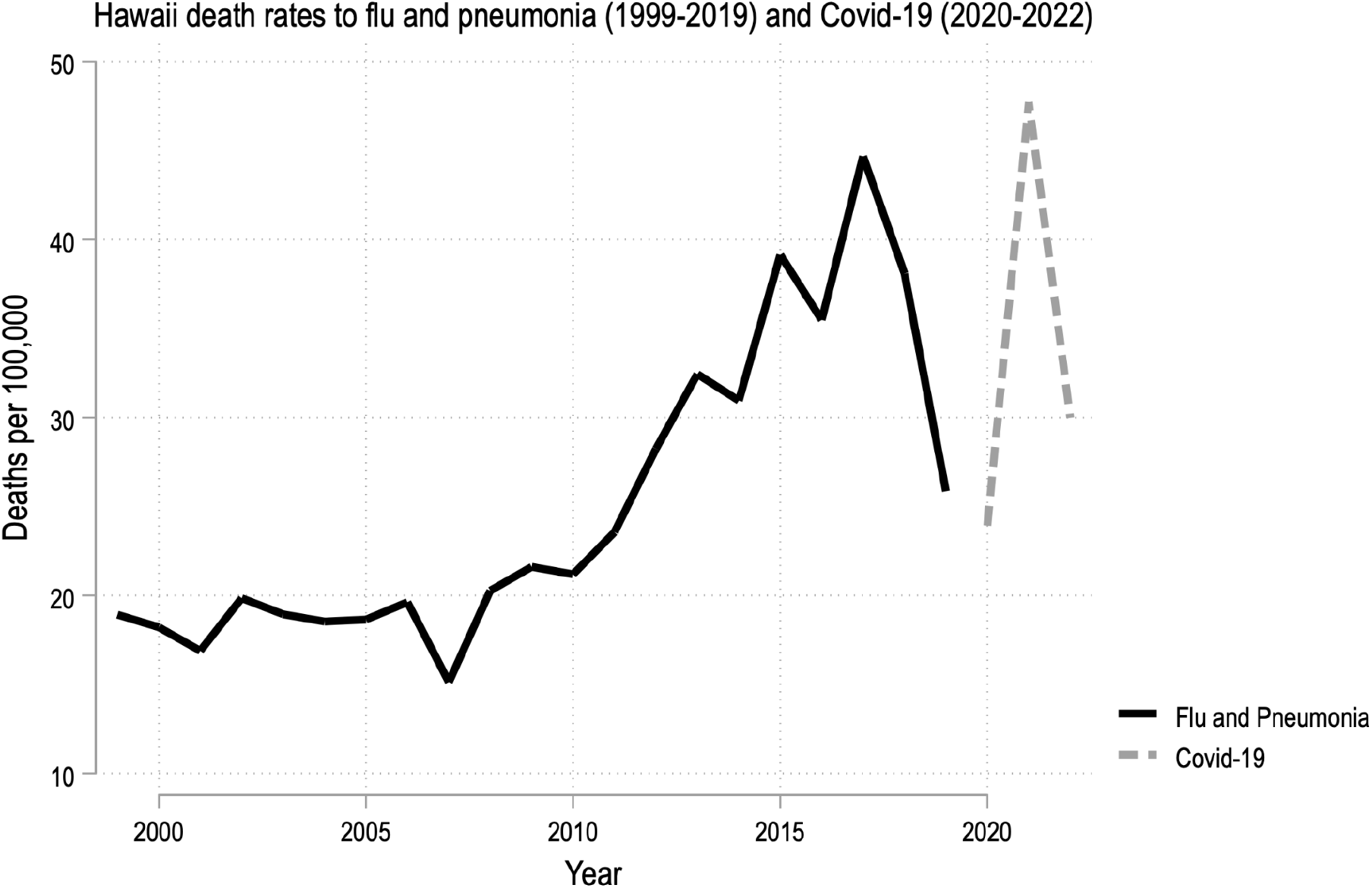
Death rates for influenza and pneumonia (1999-2019) and Covid-19 (2020-2022) in Hawaii. The Covid-19 deaths included are those in which Covid-19 is listed as the underlying cause of death, and likewise for influenza and pneumonia.

**Figure A2.**
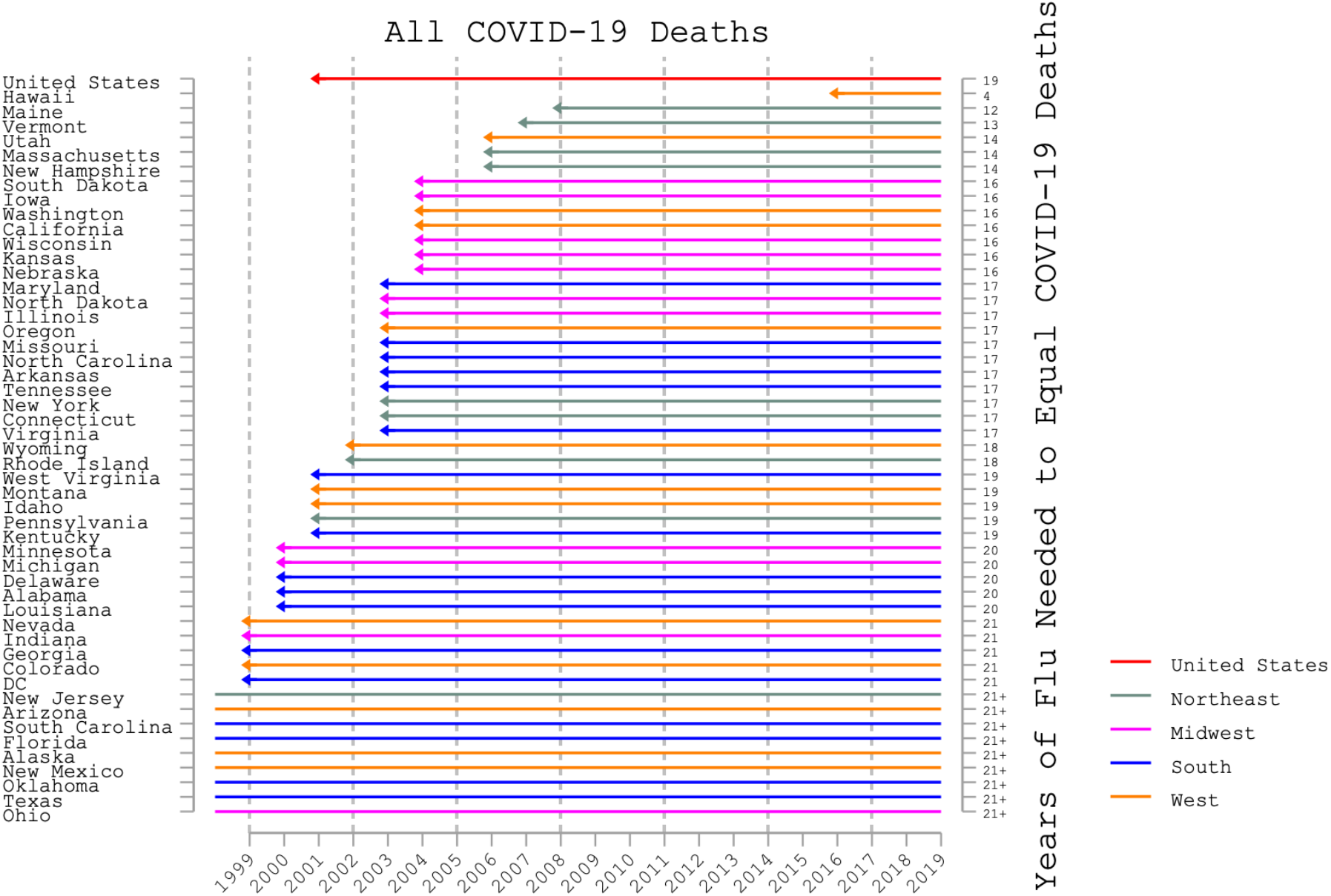
Years required for death rates to influenza and pneumonia (beginning in 2019 and counting backwards) to match the cumulative death rate to Covid-19 over a three-year period (March 2020-February 2023), for the United States as a whole (top line) and by state. In this alternative to the main graph (Fig. 1), Covid-19 deaths are those in which Covid-19 appears anywhere on the death certificate. However, influenza and pneumonia deaths are only those in which one of those causes is listed as the “underlying” cause of death, due to limitations of the historical cause of death data.

